# COVID-19 Vaccination Uptake and Self-Reported Side Effects among Healthcare Workers in Mbale City Eastern Uganda

**DOI:** 10.1101/2022.07.11.22277490

**Authors:** Gabriel Madut Akech, Andrew Marvin Kanyike, Ashah Galabuzi Nassozi, Beatrice Aguti, Ashley Winfred Nakawuki, Denis Kimbugwe, Josen Kiggundu, Robert Maiteki, Dorothy Mukyala, Felix Bongomin, Samuel Baker Obakiro, Nekaka Rebecca, Jacob Stanley Iramiot

## Abstract

**Background:** Fear of anticipated side effects has hindered the COVID-19 vaccination program globally. We report the uptake and the self-reported side effects (SEs) of the COVID-19 vaccine among Healthcare workers (HCWs) in Mbale City Eastern Uganda.

**Methods:** A cross-sectional survey of HCWs at seven different level health facilities was conducted from 6^th^ September to 7^th^ October 2021 using a structured self-administered questionnaire.

**Results:** COVID-19 vaccine had been received by 119 (69%) participants of which 79 (66%) received the two recommended doses of the AstraZeneca vaccine. Getting vaccinated was associated with working in a lower health facility (aOR= 14.1, 95% CI: 4.9 – 39.6, P=0.000), perceived minor risk of contracting COVID-19 (aOR= 12.3, 95% CI: 1.0 – 44.6, p=0.047), and agreeing that COVID-19 vaccine is protective (aOR= 16.7, 95% CI: 5.6 – 50.4, p=0.000). 97 (82%) of participants experienced side effects to at least one dose of which most were mild on both the first (n=362, 51%) and second dose (n=135, 69%). The most frequently reported side effects on the first and second dose were fever (79% *and* 20%), injection site pain (71% *and* 25%), and Fatigue (69% *and* 20%) respectively.

**Conclusions:** The majority of the HCWs in Mbale City had received at least one dose of the COVID-19 vaccine and experienced a side effect. The side effects were mostly mild on either dose thus the vaccines are generally safe.

## INTRODUCTION

Coronavirus disease-2019 (COVID-19) is a viral pneumonia caused by the novel SARS-CoV-2 which was first reported in Wuhan, China in December 2019 and continues to pose a significant threat to public health globally (1). As of 26^th^ November 2021, over 260 million cases and over 5.2 million deaths have been reported worldwide (Worldometer, 2021). In Uganda, the first case of COVID-19 was reported on 21^st^ March 2020 (3), and as of 26^th^ November 2021, over 126,965 cases and about 3,239 deaths reported (Ministry of Health Uganda, 2021).

At the moment, no single effective therapeutic agent has been approved for the treatment of COVID-19. However, several public health measures like social distancing, national-wide lockdowns, and hand-washing among others have been employed to control the pandemic(5). Despite these measures, COVID-19 remains a major public health challenge with some countries experiencing the third and fourth waves (5). Currently, vaccination against SARS CoV-2 is thought to be the best public health measure to halt the COVID-19 pandemic (6). There are currently several COVID-19 vaccine candidates with at least 8 vaccines approved for use by the World Health Organization. These include Moderna (mRNA-1273), Pfizer/BioNtech (BNT162b2), Jansen (ad26.COV2.S), Oxford/AstraZeneca (AZD1222), Covishield (Oxford/AstraZeneca formulation), Covaxin (Bharat Biotech), Sinopharm (BBIP-CorV), and Sinovac (CoronaVac) (7,8).

This first roll out of COVID-19 vaccinations was considered a land mark for overcoming the pandemic (9,10). However vaccination has long been faced with hesitancy due to public worries about side effects (SEs) and efficacy (11), which is worse with COVID-19 surrounded by conspiracies. The COVID-19 vaccines have however been reported by various clinical trials to be generally safe with limited side effects that are mild to moderate with a few severe reactions (12–15).

Healthcare workers (HCWs) are not only a high-risk group but also offer guidance on vaccine recommendations to the public which relies on them for this information (16,17). Automatic uptake of the COVID-19 vaccine by HCWs is not guaranteed as a study done in Israel, reported a high rate of COVID-19 vaccine skepticism among this group (18). Vaccination compliance relies on a personal risk–benefit perception, and broader political, religious, social, and historical factors (17).

Also, most data about side effects of the COVID-19 vaccines comes from manufacturer-funded studies in compliance with drug regulatory authorities which warrants continuous monitoring of adverse events (9,19). Ever since Uganda rolled out its vaccination program, no study has been published about the side effects of the vaccine among those that have received it. Therefore, we determined the uptake of the COVID-19 vaccine and self-reported side effects among healthcare workers in Mbale city, Eastern Uganda.

## Methods

### Study Design

This was a descriptive, cross-sectional study carried out from 6^th^ September to 7^th^ October 2021 using a quantitative approach.

### Study Setting

This study was carried out in seven health facilities in Mbale city, including six government facilities; Mbale regional referral hospital, Namatala Health centre IV, Malukhu Health Centre III, Namakwekwe Health Centre III, Busamaga Health Centre III, Mbale Municipal Health Centre II and The AIDS Support Organization (TASO) Mbale Centre of excellence a private-not-for-profit facility. Uganda’s health system has both public and private sector which has private-for-profit and private-not-for-profit facilities. The public health sector is divided into national (governed by the ministry of health) and district (governed by district health management team) based level facilities. The national level consists of national referral hospitals at the top then regional referral hospitals. The district level has the district hospitals at the helm followed by health centers IV,III, II and community extension health workers called Village health team (VHTs) serving as level I. Mbale city is located in the Eastern region of Uganda. Mbale Regional Referral Hospital is a tertiary hospital—serving the districts of Busia, Buddaka, Kibuku, Kapchorwa, Bukwa, Butaleja, Manafwa, Mbale, Pallisa, Sironko, budduda, Bukedea and Tororo(20). It’s a 470-bed capacity hospital and serves as a teaching hospital for Busitema University Faculty of Health Sciences as well as an Internship center for graduates from medical schools for a one-year internship under the supervision of qualified specialists and Consultants.

### Study Population

Healthcare workers in the selected facilities were involved in the study. A healthcare worker was considered as any person engaged in activities whose primary role is to improve the health of patients including nurses, midwives, intern doctors, medical officers, senior house officers, pharmacists, laboratory technicians and specialists in the field of medicine. Mbale Regional Referral Hospital (MRRH) had a total of 243 permanent HCWs and 57 interns on a temporary one-year contract. The other five lower health centers had a total of 120 trained health care workers of different cadres. TASO Mbale Centre of excellence had 31 full-time staff. This makes a total of about 451 health care workers in primary contact with patients within the selected health facilities in Mbale city currently.

### Inclusion and Exclusion Criteria

HCWs at selected health facilities within Mbale city who consented to participate were included and those that didn’t consent or were absent on duty were excluded.

### Sampling Procedure and Data collection

The research followed a proportional stratified random sampling technique. The study population was stratified by level of health facility each taking a proportion depending on the percentage contribution to the total required sample size. Participants were enrolled by random sampling at that particular time data was collected at the facility. Data was collected using semi-structured questionnaire that was provided by research assistants at each facility. The research assistants were trained on the unique aspects of the study via a zoom session after they had been identified. Questionnaires were administered by hardcopy after informed consent had been given by the participant.

### Data Collection tool

We used a modified questionnaire based on those that were used by other scholars. The questionnaire was semi-structured with 28 questions divided into five sections capturing socio-demographic information of participants, COVID-19 related experiences and attitudes towards COVID-19 vaccine, its uptake, and self-reported side effects among health workers that have taken the vaccine.

### Data Analysis

Data from completed questionnaires were entered in Microsoft Excel 2016, cleaned, coded and analyzed using STATA 16. Descriptive statistics, and bivariate and multivariable logistic regression analyses were performed, and results were presented as tables, charts and figures. Chi-square or Fisher’s exact test was used to assess associations between vaccine uptake and independent variables. An association with a p-value less than 0.05 was considered statistically significant.

## Results

### Baseline characteristics of the participants

One hundred seventy-two (172) participants were interviewed giving a 45% response rate. Most were female (n=95, 55%), married (n=90, 52%), with a median age of 31 (IQR: 25-39) years. Most participants were from Mbale Regional Referral Hospital (n=92, 53%). Forty-three (25%) participants were nursing officers and the minority were specialists (n=6, 3%), (Table 1).

**Table 1:**
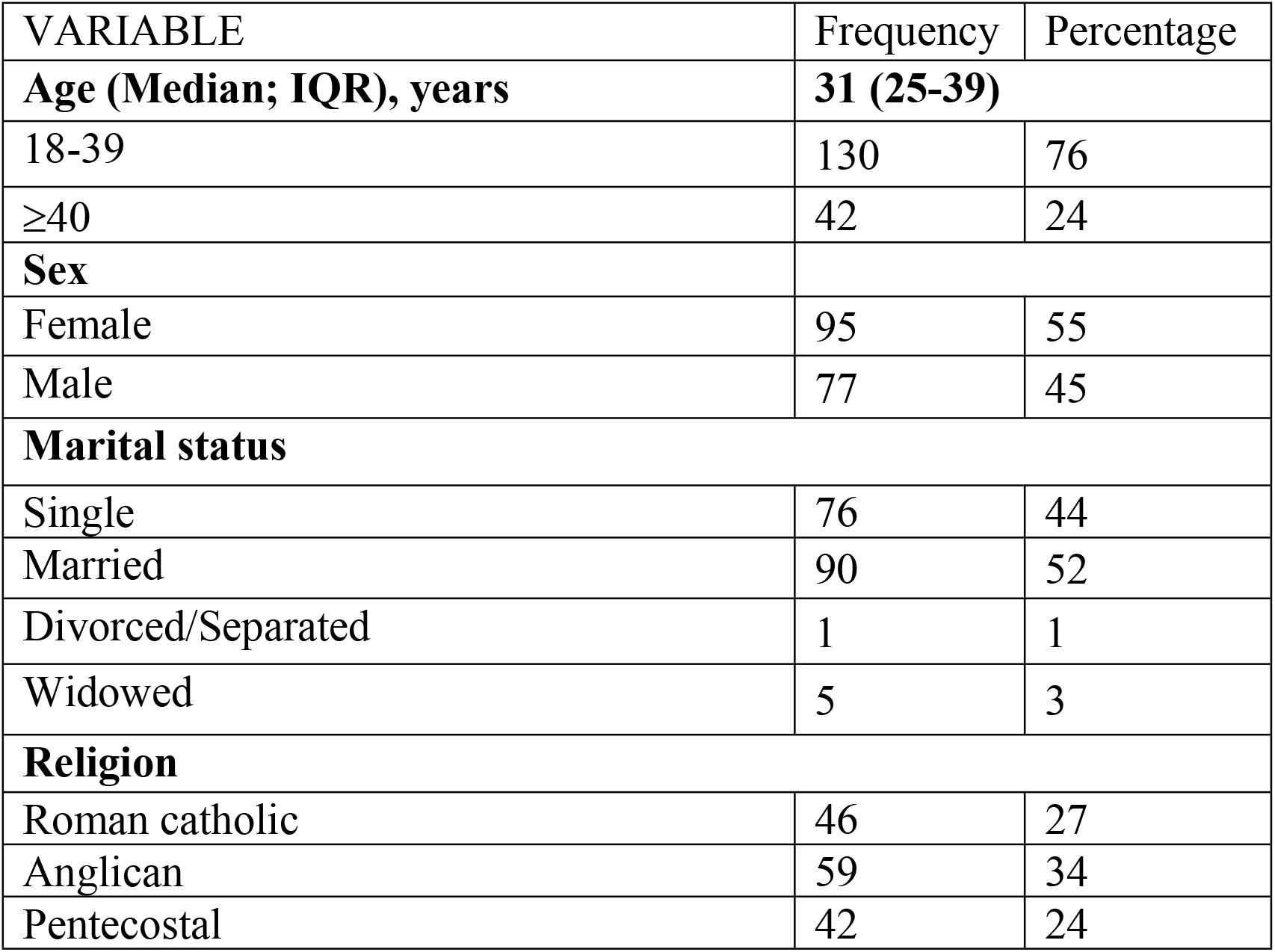
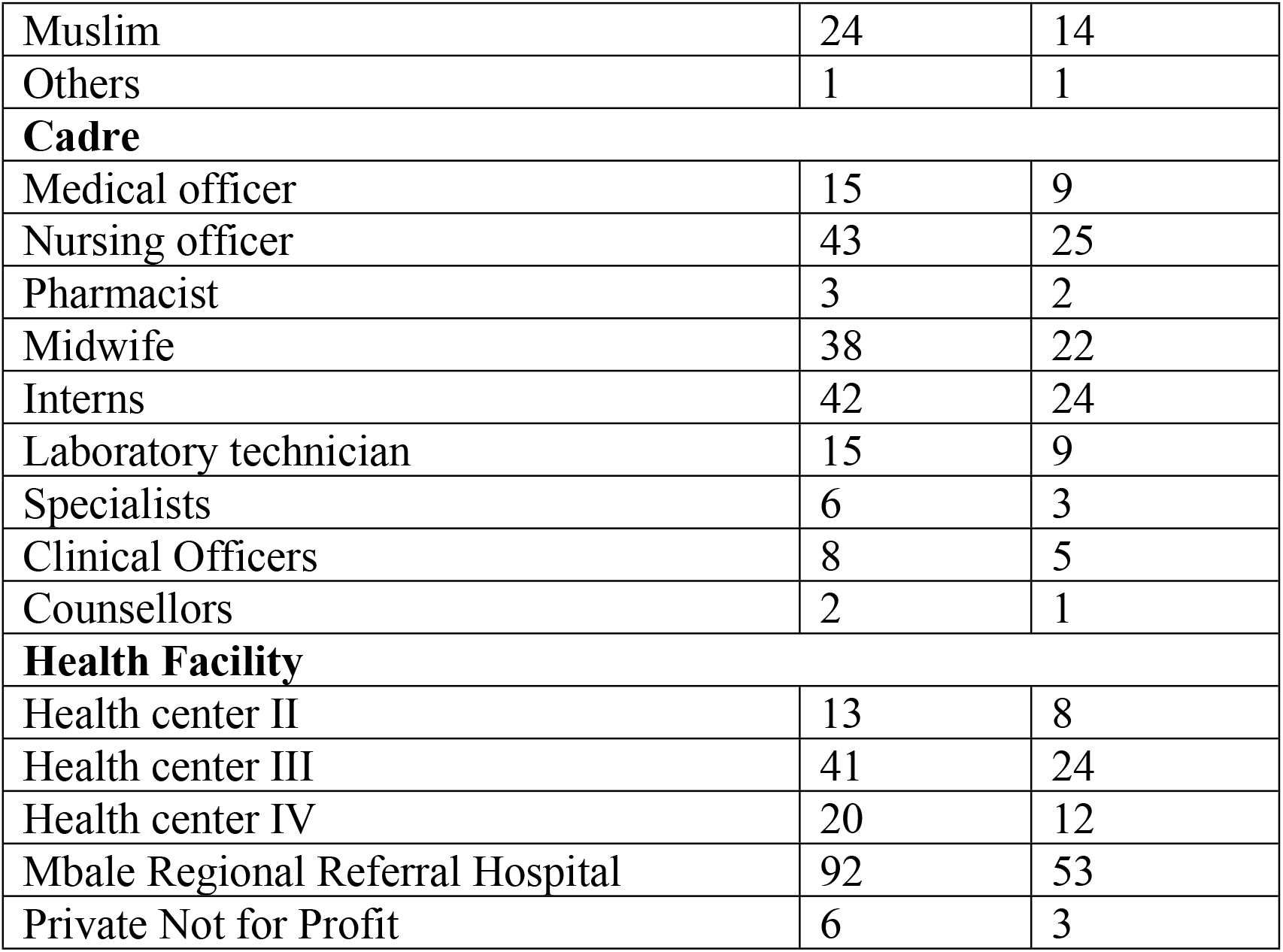
Demographic characteristic of participants (N=172)

### COVID-19 vaccine uptake among healthcare workers

The majority of the participants (n=119, 69%) had received at least one dose of the COVID-19 vaccine. Of these, 79 (66%) had received two doses. The major reason for being vaccinated was to protect oneself (n=95, 79%) and others (n=62, 52%) from COVID-19. Among unvaccinated participants, the majority (n=21, 40%) were still hesitant and not sure whether they majority will take it in future. The major reasons for not accepting to be vaccinated against COVID-19 were uncertainty about the vaccine safety (n=31, 55%), and negative information about the vaccine (n=30, 54%), (Table 2).

**Table 2:**
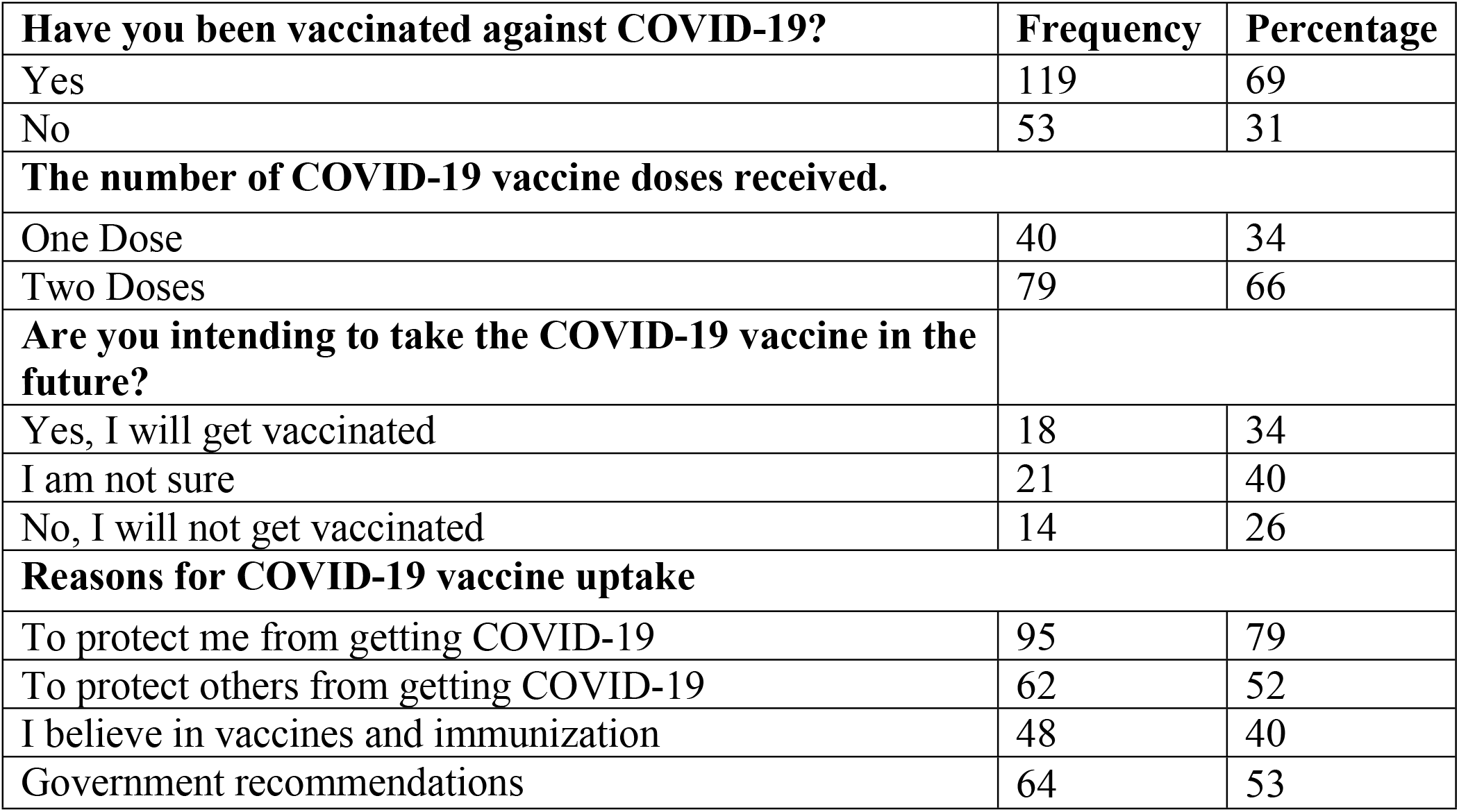
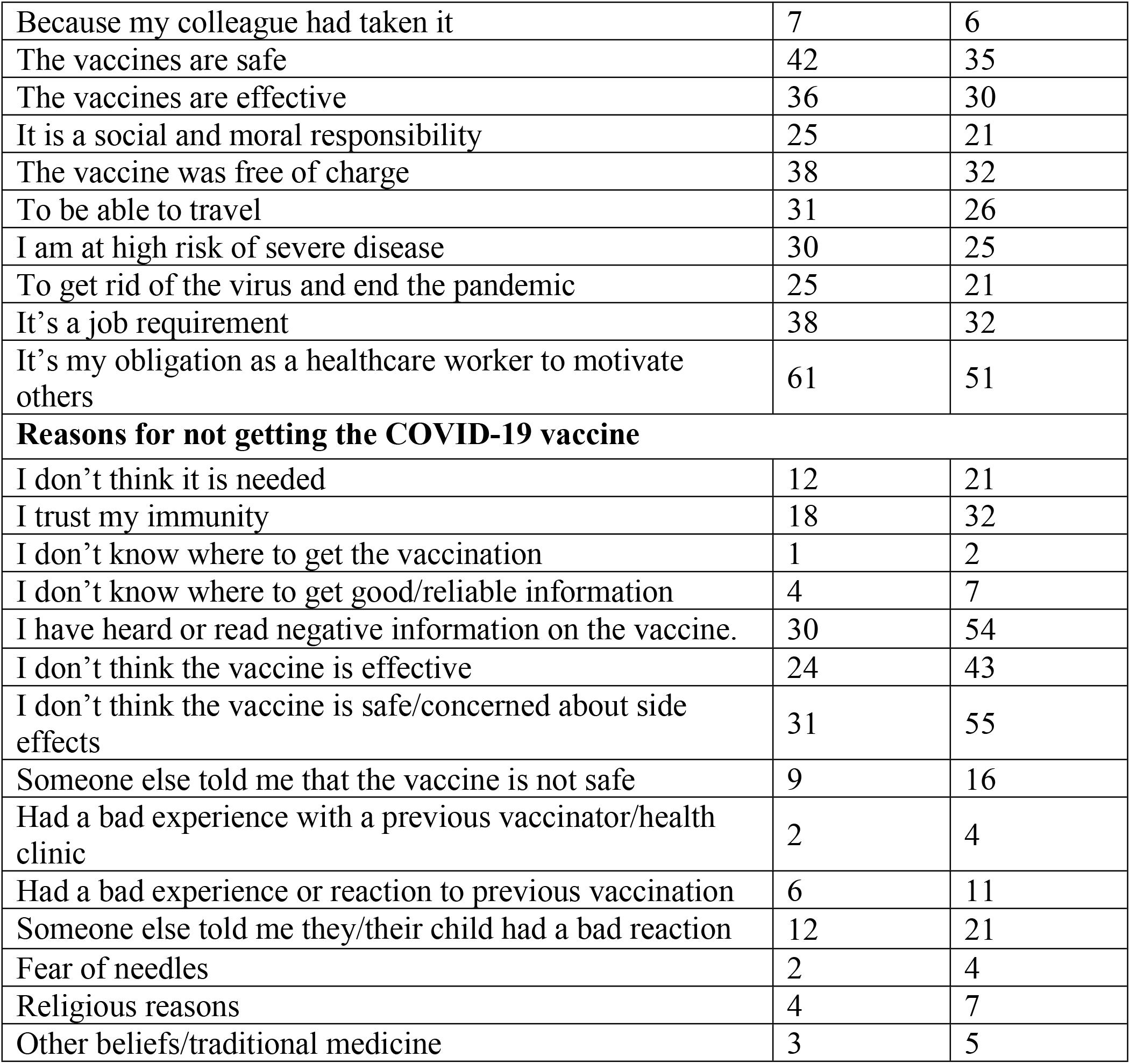
COVID-19 vaccine uptake and reasons for and against among participants.

### Factors associated with COVID-19 vaccine uptake among health workers

At bivariable analysis, age (p=0.002), marital status (p=0.019), the cadre of the staff (p<0.001), level of health facility (p<0.001), belief in the effectiveness of the COVID-19 vaccine (p<0.001) and having a chronic illness (p=0.021) were statistically significantly associated with COVID-19 vaccine uptake, Table 3.

**Table 3:**
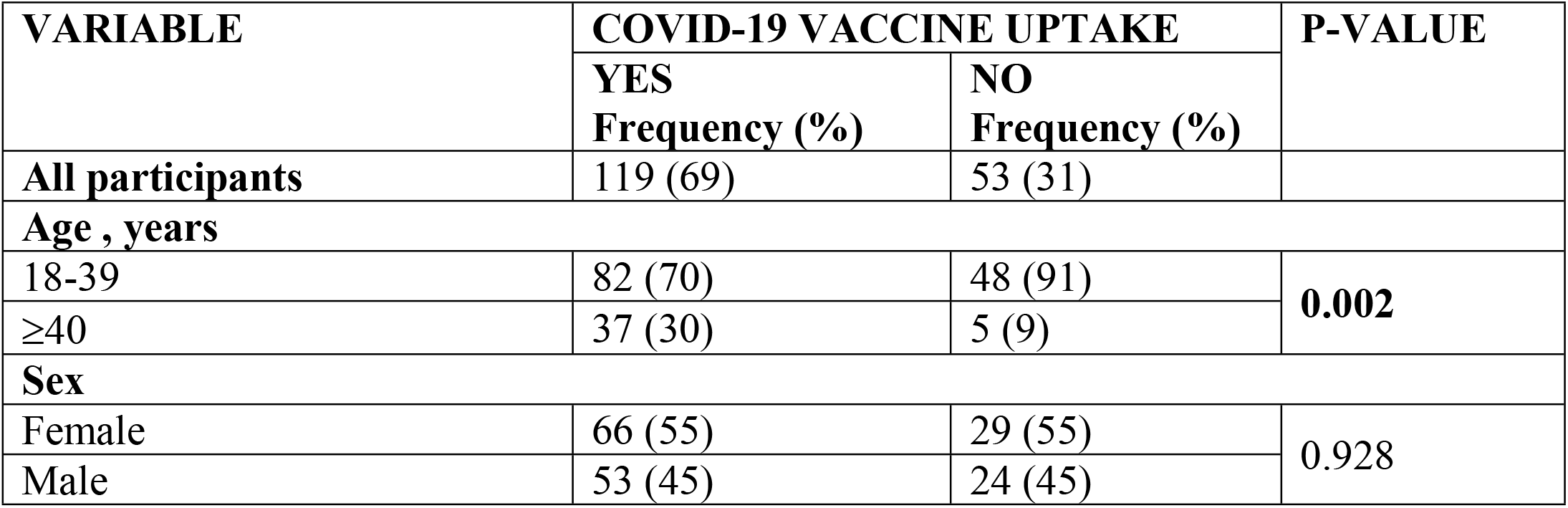
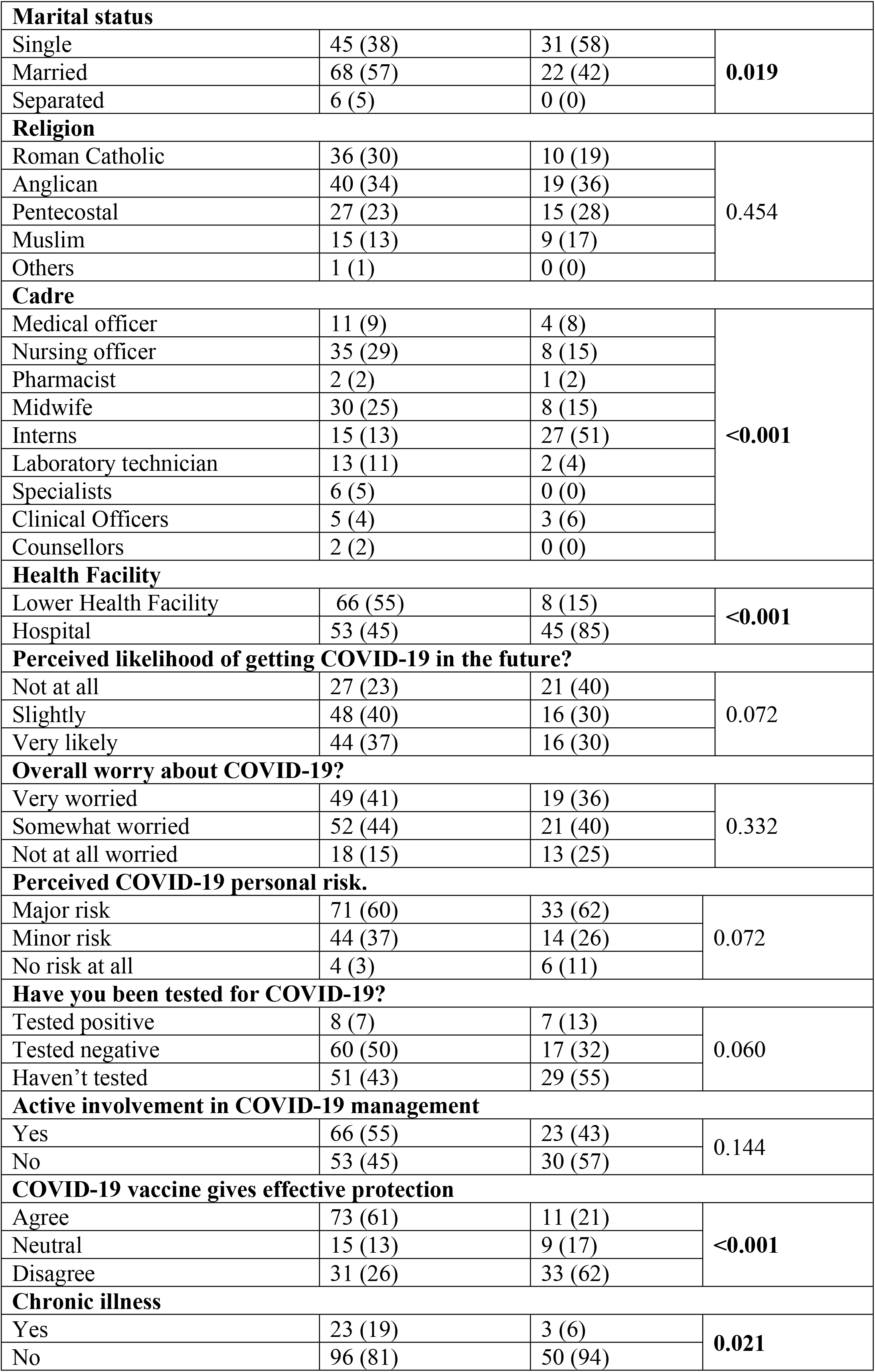
Uptake of COVID-19 vaccine and associated factors among the participants.

At multivariable analysis, factors independently associated with uptake of the COVID-19 vaccine were as follows: working in a lower health facility (aOR= 14.1, 95% CI: 4.9 – 39.6, P=0.000), individual perceived minor risk of contracting COVID-19 (aOR= 12.3, 95% CI: 1.0 – 44.6, p=0.047), and agreeing that COVID-19 vaccine is protective (aOR= 16.7, 95% CI: 5.6 – 50.4, p=0.000), (Table 4).

**Table 4:** Multivariate logistic regression for factors associated with COVID-19 vaccine uptake among participants.

| VARIABLE | OR (95% CI) | P-VALUE | AOR (95% CI) | P-VALUE |
| --- | --- | --- | --- | --- |
| **Age** |  |  |  |  |
| 18-39 | Ref |  | Ref |  |
| 40 years and above | 4.3 (1.6 – 11.8) | **0.004** | 2.7 (0.7 – 10.2) | 0.136 |
| **Marital Status** |  |  |  |  |
| Single | 0.5 (0.2 – 0.9) | **0.026** | 0.6 (0.2 – 1.6) | 0.353 |
| Separated | Ref |  | Ref |  |
| **Cadre** |  |  |  |  |
| Medical officer | 1.2 (0.2 – 6.9) | 0.856 |  |  |
| Nursing officer | 1.9 (0.4 – 8.9) | 0.428 |  |  |
| Pharmacist | 0.9 (0.1 – 13.4) | 0.913 |  |  |
| Midwife | 1.6 (0.3 – 7.7) | 0.551 |  |  |
| Interns | 0.2 (0.1 – 1.1) | 0.059 |  |  |
| Laboratory technician | 2.8 (0.4 – 20.8) | 0.318 |  |  |
| Specialists | Ref |  |  |  |
| **Health Facility** |  |  |  |  |
| Lower Health Facility | 7.0 (3.0 – 16.1) | **0.000** | 14.1 (4.9 – 39.6) | **0.000** |
| Hospital | Ref |  | Ref |  |
| **Perceived likelihood of getting COVID-19 in the future?** |  |  |  |  |
| Not at all | Ref |  | Ref |  |
| Slightly | 2.3 (1.0 – 5.2) | **0.039** | 1.8 (0.5 – 5.8) | 0.360 |
| Very likely | 2.1 (0.9 – 4.8) | 0.065 | 1.3 (0.4 – 4.4) | 0.677 |
| **Perceived COVID-19 personal risk.** |  |  |  |  |
| Major risk | 3.2 (0.9 – 12.2) | 0.084 | 8.2 (0.9 – 98.2) | 0.097 |
| Minor risk | 4.7 (1.2 – 19.1) | **0.030** | 12.3 (1.0 – 44.6) | **0.047** |
| No risk at all | Ref |  | Ref |  |
| **Have you been tested for COVID-19?** |  |  |  |  |
| Tested positive | 0.6 (0.2 – 1.9) | 0.447 |  |  |
| Tested negative | 2.0 (0.9 – 4.1) | 0.053 |  |  |
| Haven't tested | Ref |  |  |  |
| **COVID-19 vaccine gives effective protection.** |  |  |  |  |
| Agree | 7.1 (3.2 – 15.7) | **0.000** | 16.7 (5.6 – 50.4) | **0.000** |
| Disagree | 1.8 (0.7 – 4.6) | 0.242 | 1.3 (0.3 – 4.9) | 0.728 |
| Neutral | Ref |  | Ref |  |
| **Active involvement in COVID-19 management** |  |  |  |  |
| Yes | 1.6 (0.8 – 3.1) | 0.145 |  |  |
| No | Ref |  |  |  |

| Chronic illness |  |  |  |  |
| --- | --- | --- | --- | --- |
| Yes | 4.0 (1.1 – 13.9) | **0.030** | 3.1 (0.6 – 15.0) | 0.168 |
| No | Ref |  | Ref |  |

### Self-reported side-effects among covid-19 vaccine recipients

The majority of the COVID-19 vaccine recipients (n=97, 82%) experienced side-effects of at least one dose of the vaccine which lasted for a median duration of 3 (IQR: 3-7) days. All recipients (n=97, 100%) experienced at least one side effect on their first dose of vaccination and only thirty-eight (39%) on the second dose, Figure 2. Most of the side-effects were mild on both the first (n=362, 51%) and second dose (n=135, 69%). Severe side-effects were experienced more on the first (11%) than second (7%) dose (Figure 3).

**Figure 1:**
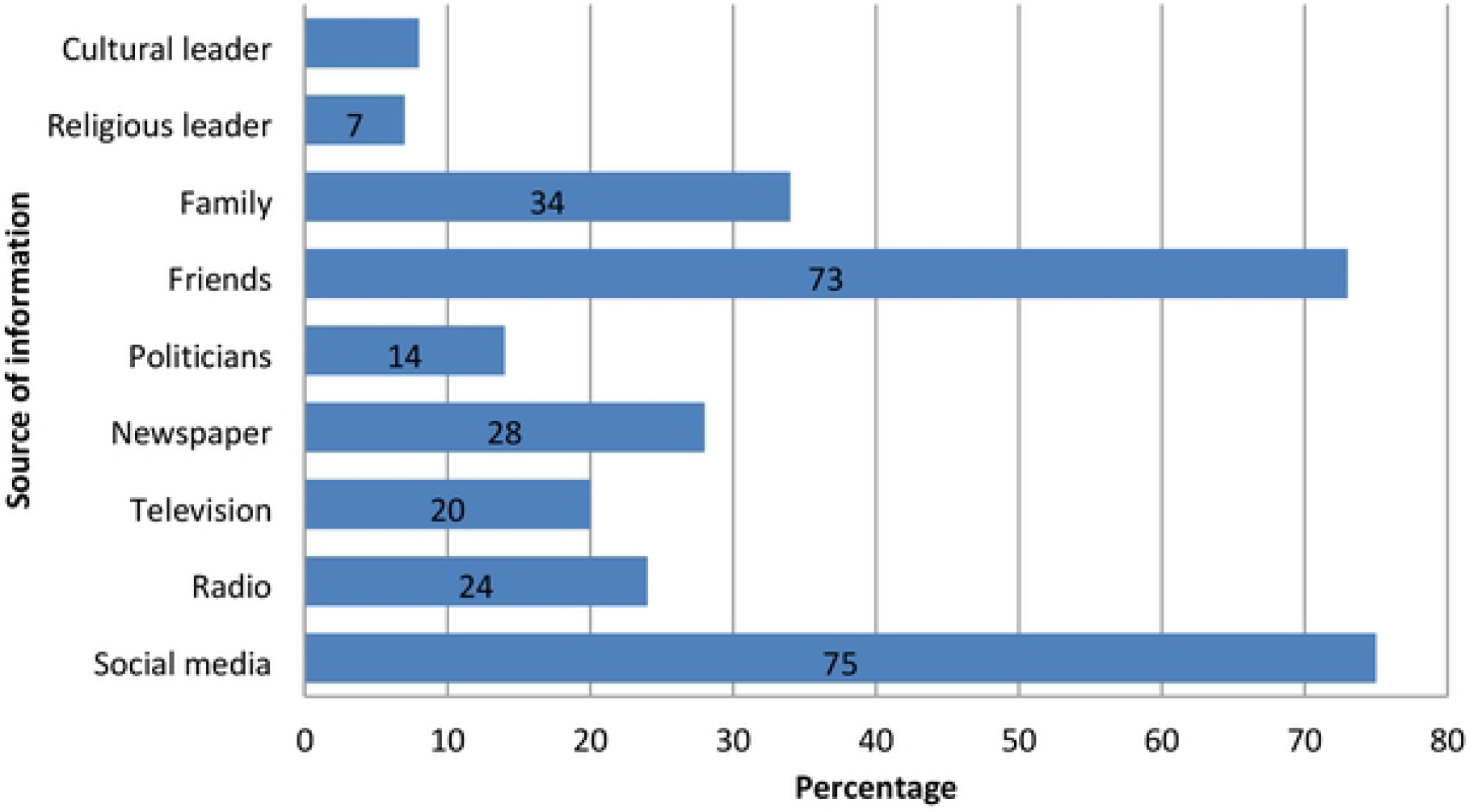
Sources of negative information about COVID-19 vaccination among participants.

**Figure 2:**
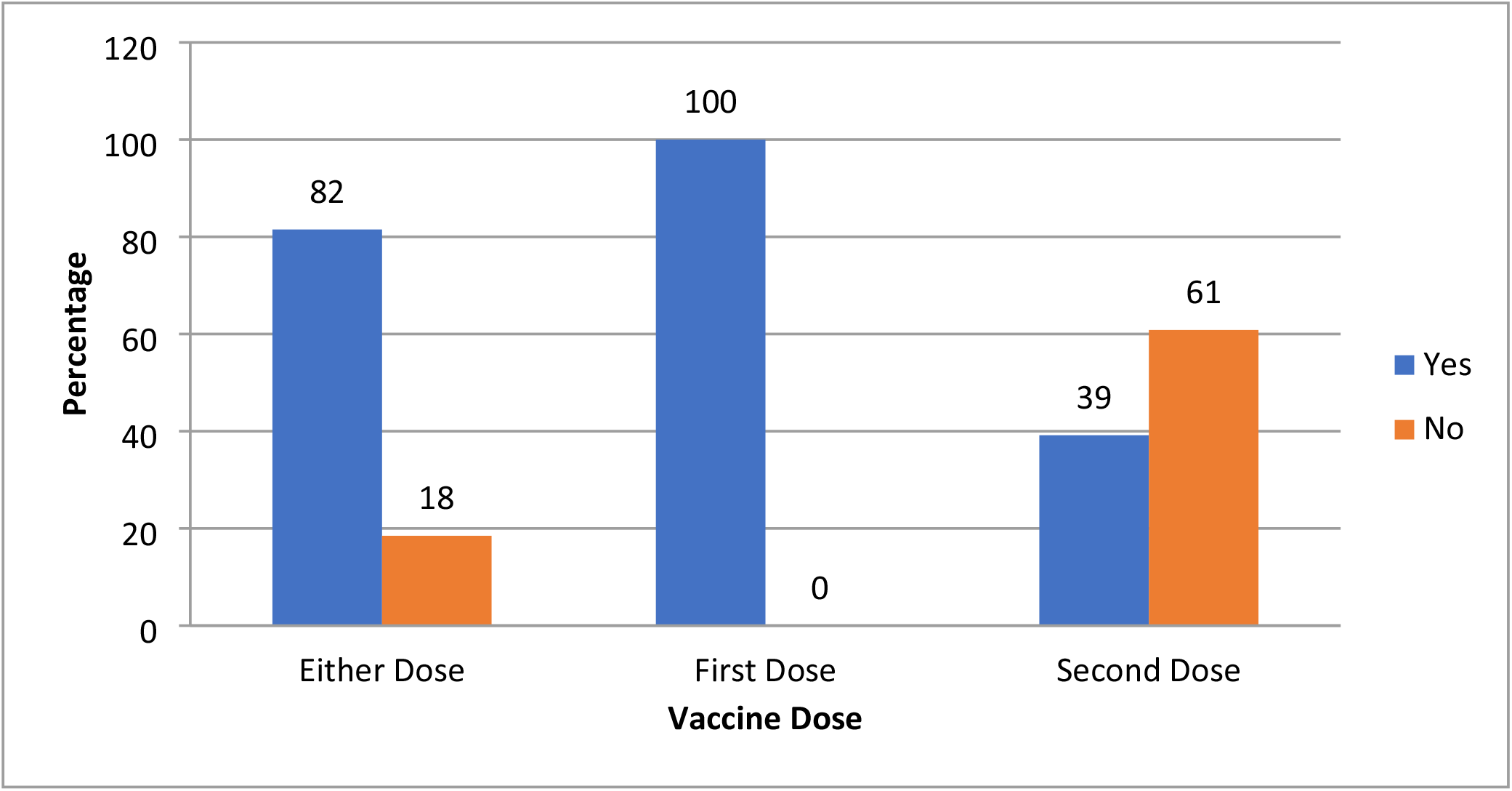
Occurrence of self-reported side-effects on the different vaccine doses.

**Figure 3:**
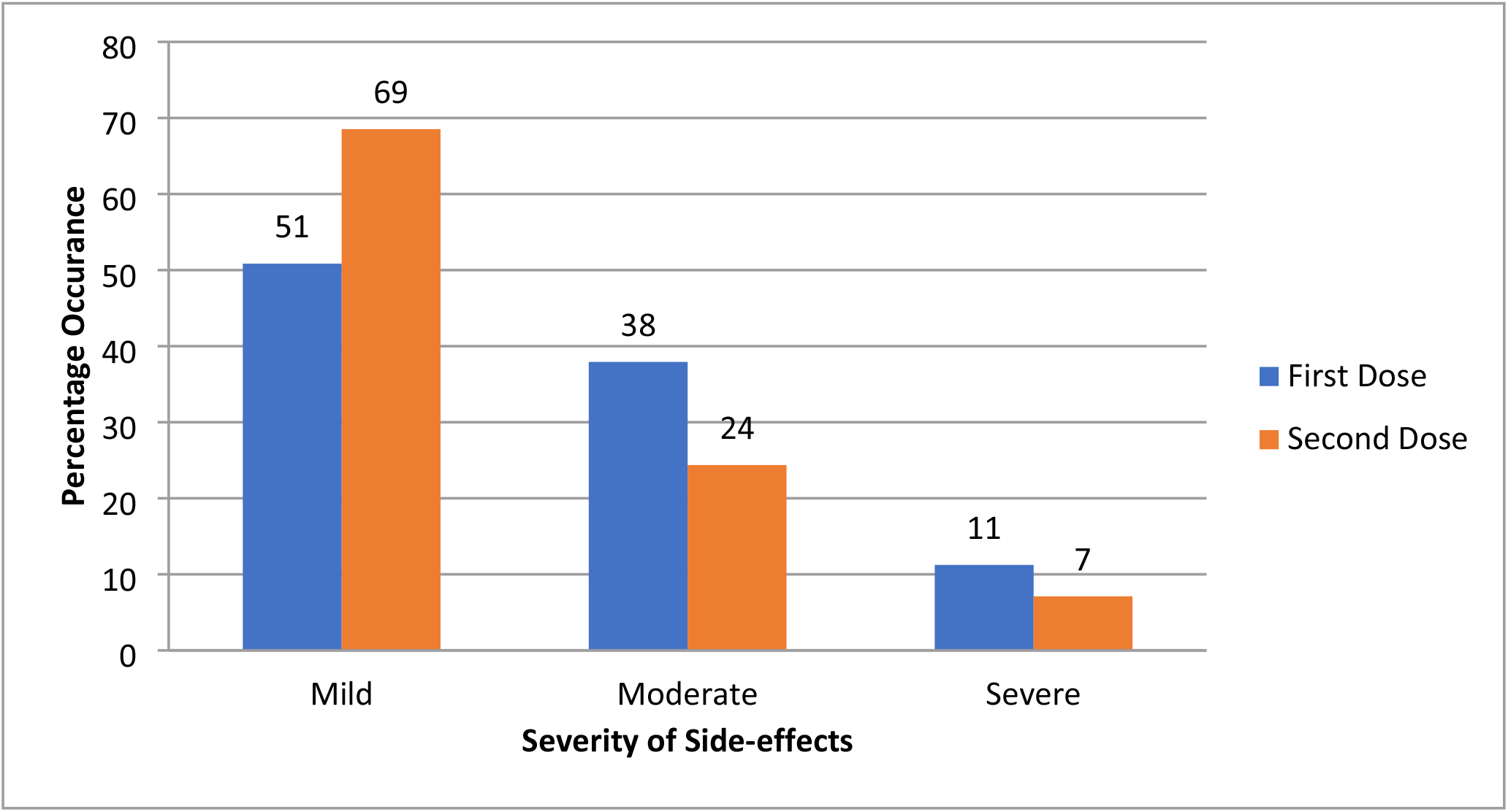
Severity of self-reported side-effects on the different vaccine doses.

The most frequently reported side-effects of the first and second dose were fever (79% and20%), injection site pain (71% *and* 25%) and Fatigue (69% *and* 20%) respectively, Figure 4. Participants who had ever tested positive for COVID-19 experienced side effects for a longer duration (median= 7, IQR: 3-11 days) compared to those who had either tested negative (median= 5, IQR: 3—5 days) or had never tested (median= 3, IQR: 3-5 days). There was no statistically significant difference in the duration of side-effects among the different age groups (Figure 5).

**Figure 4:**
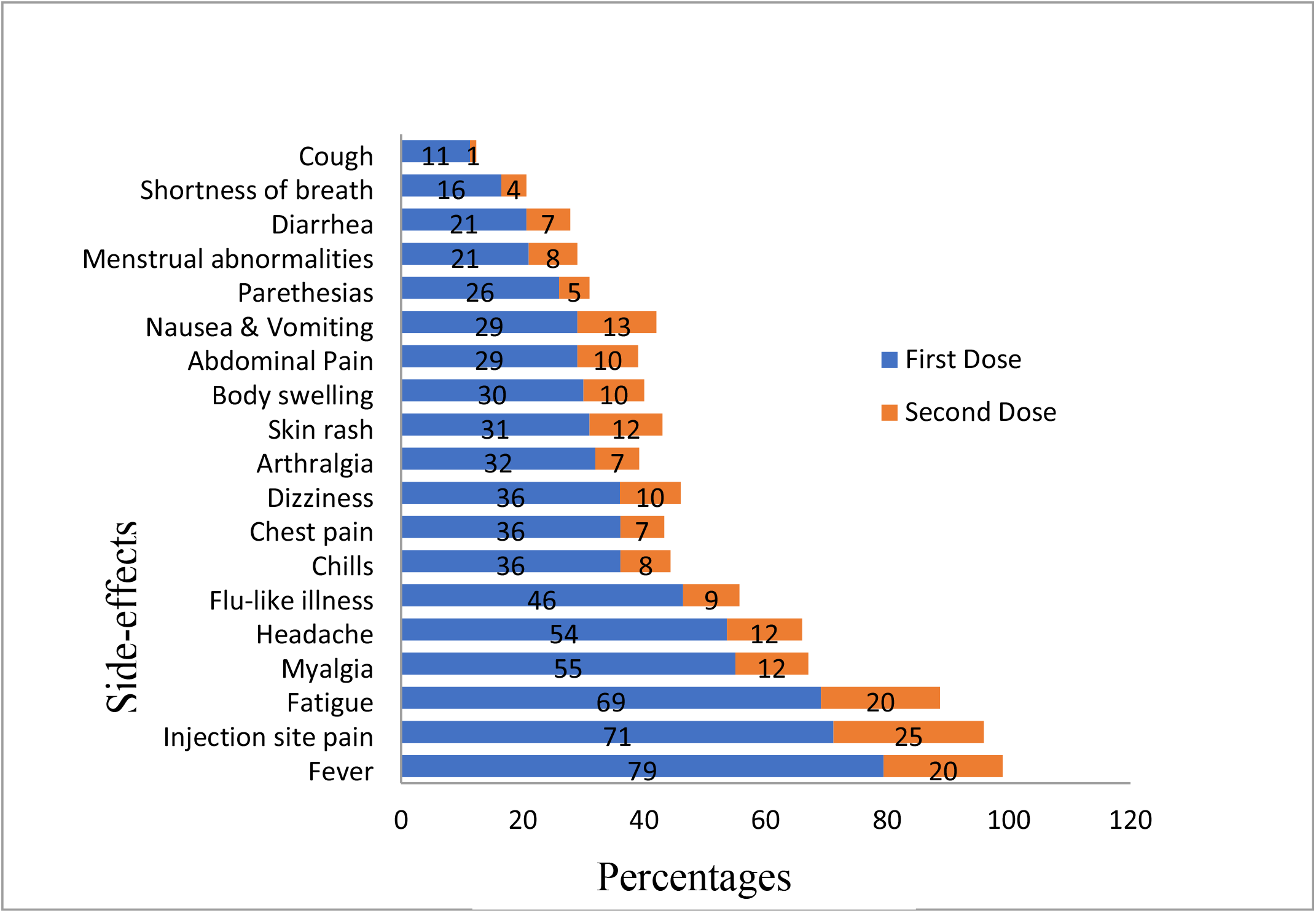
The different self-reported side effects on both COVID-19 vaccine doses.

**Figure 5:**
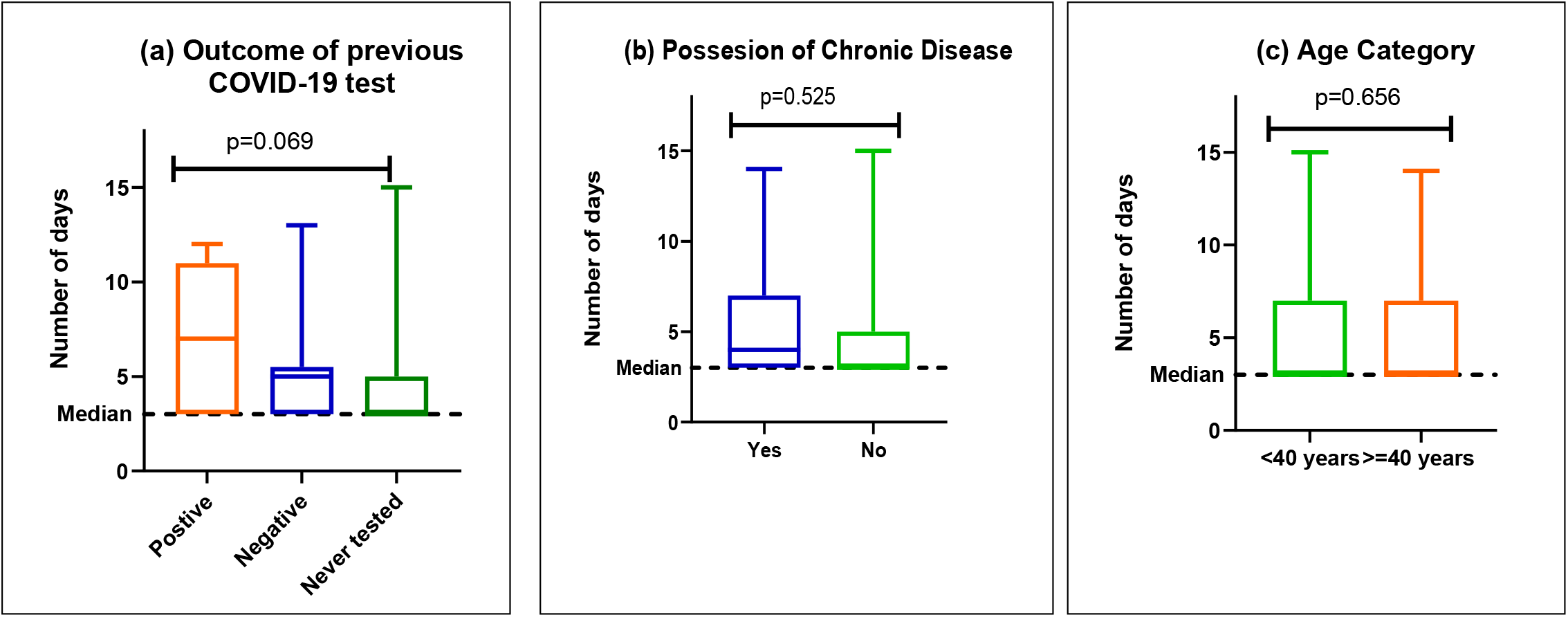
Duration of self-reported side-effects compared among (a) outcomes of previous COVID-19 test (b) possession of chronic illness and (c) age category. Median duration of side-effects is 3 days.

## DISCUSSION

Health Care Workers (HCWs) being at the frontline in the fight against the pandemic are at a high risk of contracting COVID-19 as they provide medical services to COVID-19 patients. (21). As a result several HCWs have succumbed to COVID-19, with variable morbidity and mortality reported from different countries (22). In the COVID-19 vaccination programs of most countries in low- and middle-income countries (23). HCWs have been prioritized due to the limited supply of the COVID-19 vaccine. Despite this, COVID-19 vaccine hesitancy has been reported globally, including among HCWs and the WHO reports it among the top 10 global threats to health (18).

In this study, a high proportion (79%) of HCWs had received at least one dose of the COVID-19 vaccine. Our findings are similar to the 71.1% COVID-19 vaccine uptake reported in 19 countries among healthcare workers (24). However, in. contrast, lower vaccine acceptability was reported in the U.S.A (36%) (16) and Turkey (31%) (25). Higher acceptability rates have been reported among HCWs in France (81.5%) (23), Mozambique (88.6%) (26) China (91.3%) (27), and Indonesia (95%) (28). It’s reasonable that Asian countries like China which was hit hard by the pandemic reported higher uptake of the vaccine. The participants that believed that the vaccine was effective were twenty-eight times more likely to get vaccinated. This finding is in line with another study where participants who believed that COVID-19 vaccination, is an effective way to prevent and control COVID-19 tended to accept COVID-19 vaccination as soon as possible (18). Furthermore, HCWs who perceived a major risk of contracting COVID-19 and had previously tested negative for COVID-19 took up the vaccine. Indeed in a multicenter study, Evridiki et al reported that increased risk perception towards COVID 19 was associated with the likely uptake of the COVID-19 vaccine (29). Working at a lower health facility had 25 -fold higher odds of uptake of the COVID-19 vaccine.

In this study, most of the side effects reported were mild (51%) in correspondence with the known side effects of the AstraZeneca vaccine (12–15). Most reported side effects included fever, injection site pain, and fatigue, and this is in line with a systematic review that reported the same adverse effects from various clinical trials (9,30). Participants reported more severe side effects with the first than the second dose in contrast to Mathioudakis et al who noted more side effects with the second dose (10). After the first vaccine dose, the body gets sensitized to the antigens and a severe immune reaction is expected on the booster dose. Probably most of the vaccine recipients could have been infected at the time of vaccination with sensitized immune systems that reacted severely to re-exposure of the virus since prior testing to vaccination wasn’t done. To further reinforce that, participants who had ever tested positive for COVID-19 experienced a longer duration of symptoms compared to those who tested negative.

## Limitations and Strengths

The study provides results from multiple health facilities, and information about the relatively real-world safety profiles of the AstraZeneca vaccine in Uganda. However, this study has some limitations; the sample size is relatively small and was conducted mainly in an urban setting in a single city thus the findings may not be confidently generalizable to the rest of the country. Future studies involving a larger sample size are recommended to provide a complete side-effects profile of COVID-19 vaccines among recipients in Uganda.

## Conclusion

In conclusion, our findings have shown high levels of uptake of the COVID-19 vaccine among HCWs in Mbale city. At least four-fifth of HCWs experienced at least one SE after receiving the COVID-19 vaccine. Most of the side effects were mild and the same as those seen in other commonly provided vaccines therefore the vaccines are generally safe and can be taken by the public with confidence.

## Data Availability

All relevant data are within the manuscript and its Supporting Information files.

## Consent for publication

Not applicable

## Ethical Consideration

The proposal was submitted for ethical review and approved by the Research Ethics Committee of Cure Children’s Hospital Uganda (CCHU-REC) under approval number CCHU-REC/02/2021. Additional permission was requested from the administration of the health facilities where the study was carried out and all participants provided an informed consent. The ethical principles of involvement of human research subjects as outlined in the *Nuremberg Code* and the *Declaration of Helsinki* were strictly adhered to.

## Authors’ contributions

This study was carried out with considerable contributions from all authors. Author FB, AMK, AGN, BA, AWN, GMA, RM, JSI and SBO, conceptualized and design the study protocol. Author AMK, GMA, AGN, BA, AWN, DK, JK, RM, and DM, participated in data collection. AMK, FB, GMA and JSI analyzed the data. Author GMA and AMK drafted the first manuscript, NK and JSI conducted critical reviews. All authors reviewed and approved the final manuscript.

## Authors’ Details

^1^Faculty of Health Sciences, Busitema University, Mbale, Uganda, ^2^Mbale Regional Referral Hospital, Ministry of Health, Uganda, ^3^Ministry of Health/AIDS Control Program, Kampala, Uganda, ^4^Department of Medical Microbiology and Immunology, Faculty of Medicine, Gulu University, Gulu, Uganda, ^5^Department of Pharmacology and Therapeutics, Faculty of Health Sciences, Busitema University, Mbale, Uganda, ^6^Department of Public Health, Faculty of Health Sciences, Busitema University, Mbale, Uganda, ^7^Department of Microbiology and Immunology, Faculty of Health Sciences, Busitema University, Mbale, Uganda

